# Multivariate Analysis for the Influence of Ejection Fraction Value on Propofol Anesthesia Induction Requirement and its Pharmacodynamic Properties in Cardiopulmonary Bypass Surgery (Observational Study based on the Clinical Outcomes)

**DOI:** 10.1101/2022.11.28.22282821

**Authors:** Dias Permeisari

**Affiliations:** Department of Clinical Pharmacy, Faculty of Pharmacy, Universitas Airlangga, Indonesia

## Abstract

**Background:** The majority of propofol utilization as an induction anesthetic in cardiac surgery, particularly in cardiopulmonary bypass surgery led to several risks to the patient. The most common risk is dropped mean arterial pressure, even with the high risk of cardiac arrest.

**Objective:** Determining the influences of ejection fraction value on the amount of propofol requirement as an induction agent based on the patient’s primary outcome (BIS spectral index) and the secondary outcomes (mean arterial pressure and heart rate)

**Design:** Prospective study, analytical observational with multivariate linear regression analysis, and multicenter study

**Setting:** 2 hospitals, including 1 teaching hospital and 1 private hospital

**Patients:** all patients who underwent cardiopulmonary bypass surgery and are eligible for the inclusion criteria

**Measurements:** Doses of Propofol as an anesthesia induction, mean arterial pressure (MAP) prior to surgery, heart rate (HR) prior to surgery, BIS Spectral Index prior to surgery, MAP after induction, HR after induction, and BIS Spectral Index after induction

**Results:** These data were analyzed using MATLAB R2022a software to obtain R^2^ (determining the effect size or influences) and p-value for each condition of ejection fraction value and the clinical responses. The data of this observational study is divided into six groups : 1. the effect size of ejection fraction value < 50% on BIS index obtained R^2^ 0.9231 and p-value 0.88, 2. the effect size of ejection fraction value ≥ 50% on BIS index obtained R^2^ 0.7794 and p-value 0.01, 3. the effect size of ejection fraction value < 50% on mean arterial pressure obtained R^2^ 0.00024 and p-value 0.97, 4. The effect size of ejection fraction value ≥ 50% on mean arterial pressure obtained R^2^ 0.0786 and p-value 0.005, 5. The effect size of ejection fraction value < 50% on heart rate obtained R^2^ 0.3992 and p-value 0.06, 6. The effect size of ejection fraction value ≥ 50% on heart rate obtained R^2^ 0.1757 and p-value 7.0776e-04.

**Conclusions:** Propofol extremely impacts BIS index value compared to the patient’s mean arterial pressure or heart rate at the induction doses of propofol in general anesthesia for patients with a reduced ejection fraction

## INTRODUCTION

Propofol is one of the anesthetic class of drugs given intravenously as an induction agent in general anesthesia which can be given to patients with a minimum age of more than three years, as well as maintenance in surgery in patients with a minimum age of more than two months, although it is also commonly used in patients in the ICU for sedation ^(1)^. As an anesthetic agent, propofol acts as an inhibitory neurotransmitter GABA to keep channels open resulting in an increased chloride conductance through neurons and hyperpolarization of cell membranes ^(2)^. The use of propofol in cardiac surgery will cause significant hypotensive and bradycardic effects, in addition to the side effects of respiratory depression, as well as very unstable hemodynamics, thus the titration is needed in administering propofol ^(3)^. The incidence of cardiac arrest caused by respiratory arrest related to anesthesia is mostly caused by the use of general anesthetic techniques which reaches almost 13% and can cause patient death ^(4)^.

Patients undergoing CABG surgery are patients who have a history of coronary artery disease (CAD) with triple vessel disease (TVD) in which there is the narrowing of all three coronary epicardial arteries (left anterior descending : LAD, left circumflex artery : LCX, right coronary artery : RCA) which reached ≥ 50% ^(5)^ and these patients generally also have several co-existing diseases such as hypertension, diabetes mellitus, and previous stroke ^(6)^.

The use of propofol as an induction agent in general anesthesia is closely related to pharmacokinetic and pharmacodynamic applications. Some of the pharmacokinetic implications in the clinical setting are aimed to minimize negative side effects and obtaining the desired therapeutic results. Estimation of the proper drug concentration is strongly influenced by pharmacokinetic calculations to determine the frequency of drug administration, drug dosage forms, and the patient’s pathophysiological condition ^(7)^. As in heart failure (HF) patients, several physiological parameters can affect the pharmacokinetic and pharmacodynamic profiles of drugs, further studies are needed to compare PK/PD profiles in healthy individuals and patients with heart failure ^(8)^.

The primary outcome in this observational study was the BIS spectral index value measured after the administration of propofol as an anesthetic induction in general anesthesia. BIS spectral index is a method used to determine the level of consciousness of an anesthetized patient. The BIS index value is always maintained in the range of 40-60 after administration of anesthesia, this aims to avoid awareness in anesthetized patients and CNS depression due to excessive anesthetic doses ^(9)^. According to one study, BIS spectral responded best to propofol, compared to sevoflurane and midazolam ^(10)^.

### Previous Studies and New Approaching

Several studies have been carried out previously were the combination of propofol with other anesthetic drugs and related it to the resulting clinical response, but there are still rare studies that have been carried out previously which aim to link the propofol dose requirement with the patient’s pathophysiological condition. Meanwhile, based on the theory described in the previous section, it is possible that the PK/PD profile of drugs may experience changes in patients with heart failure conditions. In this study, a new approach was taken to consider the ejection fraction value of an individual and classify it into two categories, namely normal and mid-reduced ejection fraction, then relate it to the propofol dose requirement and the clinical response produced by patients with different ejection fraction values.

### Left Ventricular Ejection Fraction Value

Left ventricular ejection fraction (LVEF) describes the function of left ventricular systole, where LVEF is a volume of blood ejected during systole and is also related to the volume of blood in the ventricle during end diastole (EDV) ^(11)^. In HF patients, the LVEF value is one of the determinants of patient therapy and the EF value can be classified into three, namely : normal ejection fraction (≥ 50%), mid-reduced ejection fraction (≤ 50% and ≥ 35%), and low ejection fraction (≤ 35%) ^(12–13)^.

## METHODS

The subjects involved in this study were all patients who underwent CABG surgery at Premier Hospital and Dr.Soetomo General Hospital Surabaya, Indonesia, who met the study inclusion criteria, those criteria are receiving propofol as induction of anesthesia in general anesthesia with the order of drug procedures given, those are midazolam – fentanyl – propofol – rocuronium, and adult patient (≥ 18 years and ≤ 65 years). While some patients who were excluded by the observer in this study were patients who had abnormal heart rate values before undergoing surgery, patients with a severe addiction to alcohol and smoking, patients who had a history of end-stage renal disease (ESRD), and chronic liver disorders. The induction of anesthesia was carried out by different anesthesiologists, but the anesthetic procedure performed by each doctor was the same, even though the study was conducted in two different hospitals. The procedure for administering general anesthesia is midazolam (0.05 – 0.1 mg/kg) given for the first time, followed by administration of fentanyl 2 micrograms/kg, propofol given according to the needs of the patient starting with administration of the smallest dose of propofol, namely 0.5 mg/ml and given intermittently, titration or gradually, and the last is the administration of a relaxant agent, namely rocuronium (1 mg/kg). Observation of the BIS index, MAP, and HR values was carried out for 15 minutes after the patient entered the operating room and installed tools to measure the BIS index, MAP, and HR. The first observation of the BIS index, MAP, and HR before the patient was given anesthesia induction was carried out for 3 minutes. The second observation was made after the patient was started to be given induction of midazolam, fentanyl, propofol, and rocuronium. Observation of the BIS index, MAP, and HR was carried out after the administration of each induction agent. The average time required by each anesthesiologist in giving each induction agent from the first to the second, and up to the fourth agent is about 10-15 seconds respectively, as of the observation of the clinical response after administration of each induction agent was carried out for 10 seconds.

## RESULTS

The characteristics of all patients who took part in this study are listed in table 1. The total number of patients who took part in this study from May 25, 2022, to July 24, 2022, was 15 subjects of which 6 subjects were excluded from this study, so there were only 9 subjects who met the inclusion criteria in this study. The nine subjects consisted of 3 subjects with mid-reduced ejection fraction values and 6 subjects with normal ejection fraction values. Some of the excluded subjects were due to the administration of anesthesia that was not in accordance with the statutory procedure, the patient died when he was about to be operated on, or the patient was canceled from surgery or the postponement of the operation. This study has been approved by The Research Ethics Committee of both hospitals. The procedure for administering anesthesia by the cardiac anesthesiologist to each subject is the same, both for subjects with normal ejection fraction values and for subjects with mid-reduced ejection fraction values.

**Table 1.**
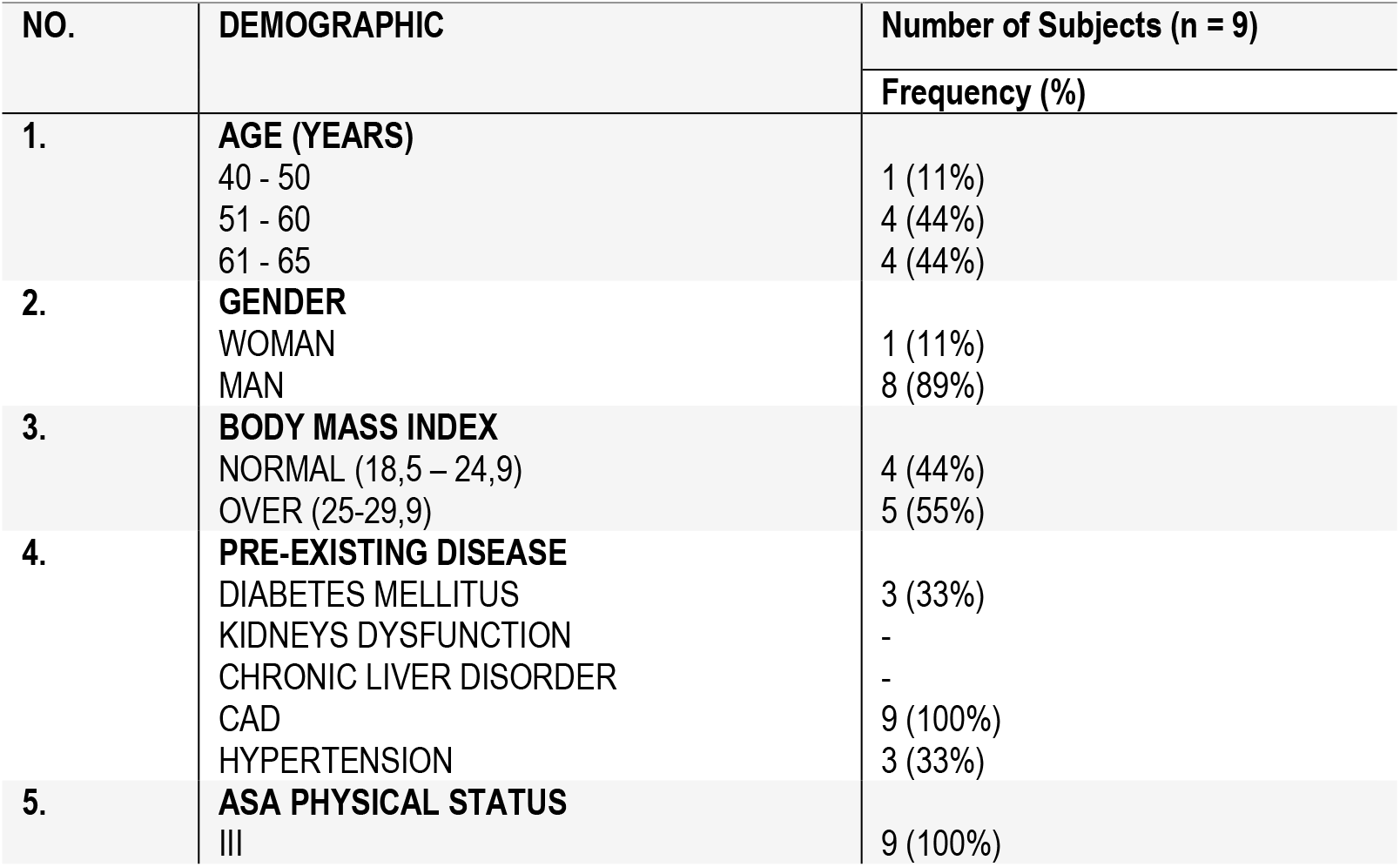
Demographic Characteristics of Subjects.

Observation of the clinical response after induction administration is divided into 6 conditions as shown in Figures 1–6. The data analysis was performed by multivariate linear regression using MATLAB R2022a software to obtain the effect size or the influence of ejection fraction value on the dose of propofol requirement and the resulting clinical response (R^2^), the norm of residuals for each of these conditions, and the p-value for each different conditions. Each patient’s ejection fraction value, BIS index value, MAP, and HR before and after induction administration are listed in table 2.

**Table 2.** Ejection Fraction Value and Clinical Responses Before and After Propofol Administration.

| No. Patient | Total Doses of Propofol (mg) | Ejection Fraction value (%) | BIS index value |  | Mean Arterial Pressure |  | Heart Rate |  |
| --- | --- | --- | --- | --- | --- | --- | --- | --- |
|  |  |  | Before | After | Before | After | Before | After |
| 1 | 100 | 45 | 95 ± 3 | 25 ± 3 | 72 ± 2 | 59 ± 5 | 70 ± 2 | 63 ± 3 |
| 2 | 30 | 32 | 80 ± 7 | 55 ± 5 | 68 ± 5 | 68 ± 5 | 70 ± 3 | 70 ± 3 |
| 3 | 50 | 60 | 99 | 70 ± 5 | 70 ± 8 | 70 ± 8 | 77 ± 5 | 77 ± 5 |
| 4 | 70 | 62 | 95 ± 5 | 63 ± 2 | 95 ± 5 | 61 ± 2 | 70 ± 5 | 60 ± 3 |
| 5 | 200 | 63 | 94 ± 3 | 44 ± 4 | 98 ± 2 | 78 ± 5 | 70 ± 2 | 65 ± 3 |
| 6 | 100 | 65 | 100 | 70 ± 4 | 76 ± 3 | 57 ± 3 | 78 ± 2 | 72 ± 3 |
| 7 | 60 | 68 | 90 ± 5 | 70 ± 2 | 78 ± 3 | 78 ± 3 | 75 ± 2 | 75 ± 2 |
| 8 | 100 | 68 | 97 | 60 ± 3 | 78 ± 2 | 78 ± 2 | 75 ± 2 | 65 ± 5 |
| 9 | 80 | 46 | 100 | $47 \pm 2$ | $75 \pm 2$ | $93 \pm 3$ | $74 \pm 2$ | $67 \pm 3$ |

**Figure 1.**
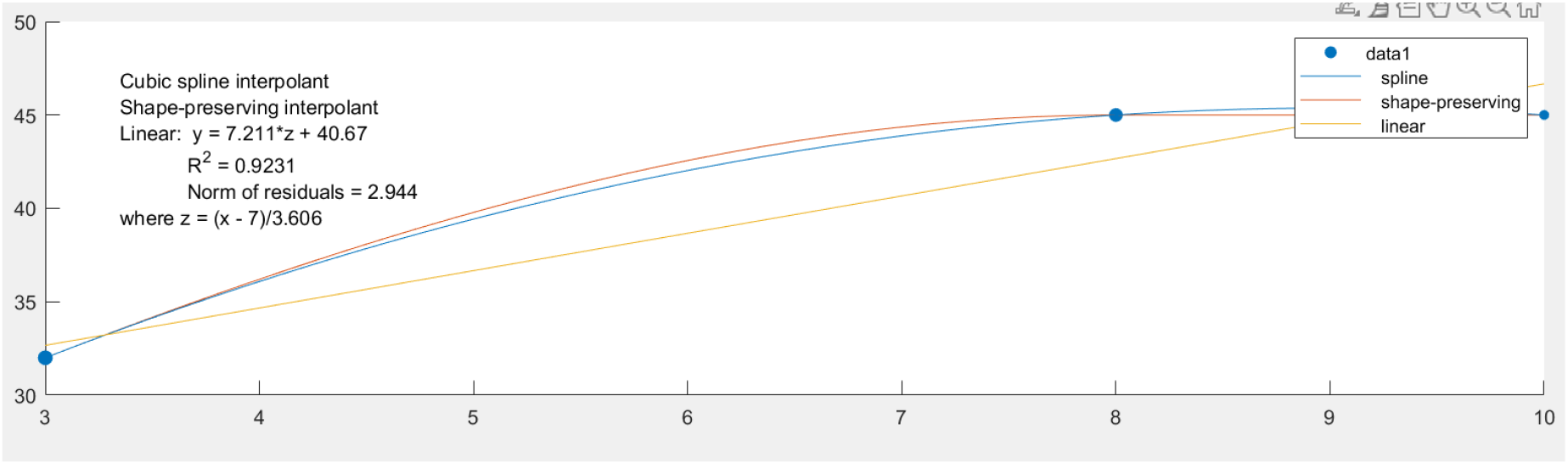
The Effect Size of Ejection Fraction Value < 50% on the BIS Index Value. In figure 1, the x-axis shows the dose of propofol in mg/ml and the y-axis shows the BIS index value, the number of subjects included in the ejection fraction <50% for measuring the response to the BIS index value is 3 subjects with an analysis result of p-value = 0.88. Where the BIS index was measured before the patient was given an induction, then another observation was made of the BIS index value after the patient was given midazolam, fentanyl, propofol, and rocuronium. Before the patient is given an induction, the BIS index value is on average 100 which indicates that the patient is at a full level of consciousness. When the anesthetist finished giving midazolam to the patient, the patient’s BIS index value still showed 100, this was the same as after giving fentanyl to the patient, the BIS index still showed However, after the anesthetist gave propofol induction, the patient’s BIS index value showed a decrease significant in this study group.

**Figure 2.**
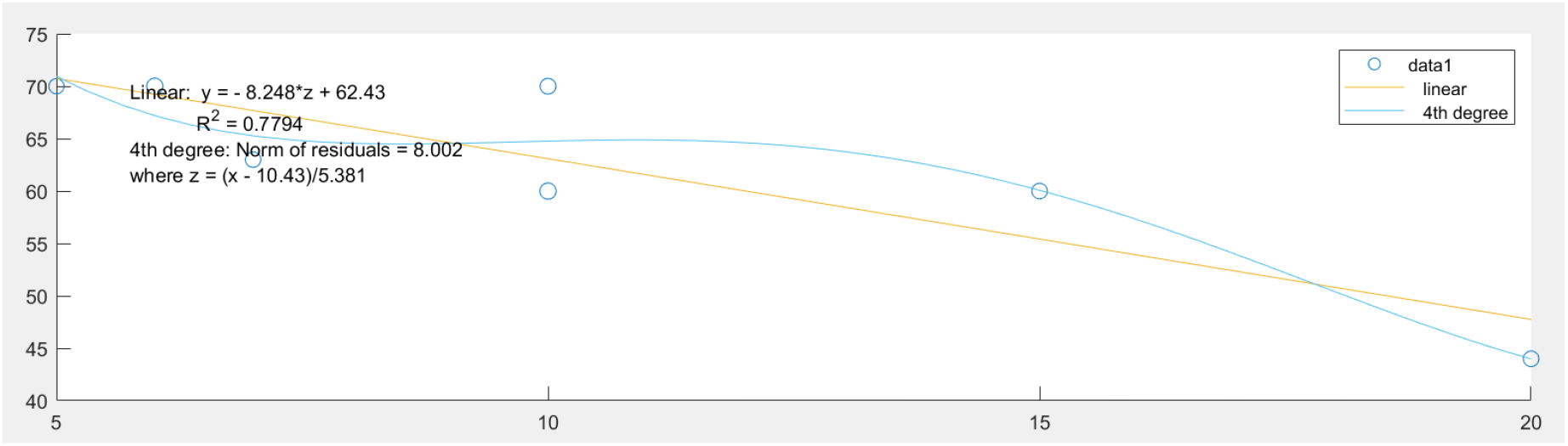
The Effect Size of Ejection Fraction Value ≥ 50% on the BIS Index Value. In figure 2, the x-axis shows the dose of propofol given in mg/ml and the y-axis shows the BIS index value, the number of subjects included in the category of ejection fraction value ≥ 50% is 6 subjects, with a p-value = 0.01. Observation of the BIS index value was carried out starting before the patient received anesthetic induction and after the induction agent was given. Based on the results of this observational study, it shows that the dose of propofol requirement to be able to produce a BIS index value in the range of 40-60 is greater than the dose of propofol in patients with a reduced ejection fraction. Based on the R^2^ produced in this study group, the dose of propofol-induced can provide a considerable influence on changes in the value of the BIS index.

**Figure 3.**
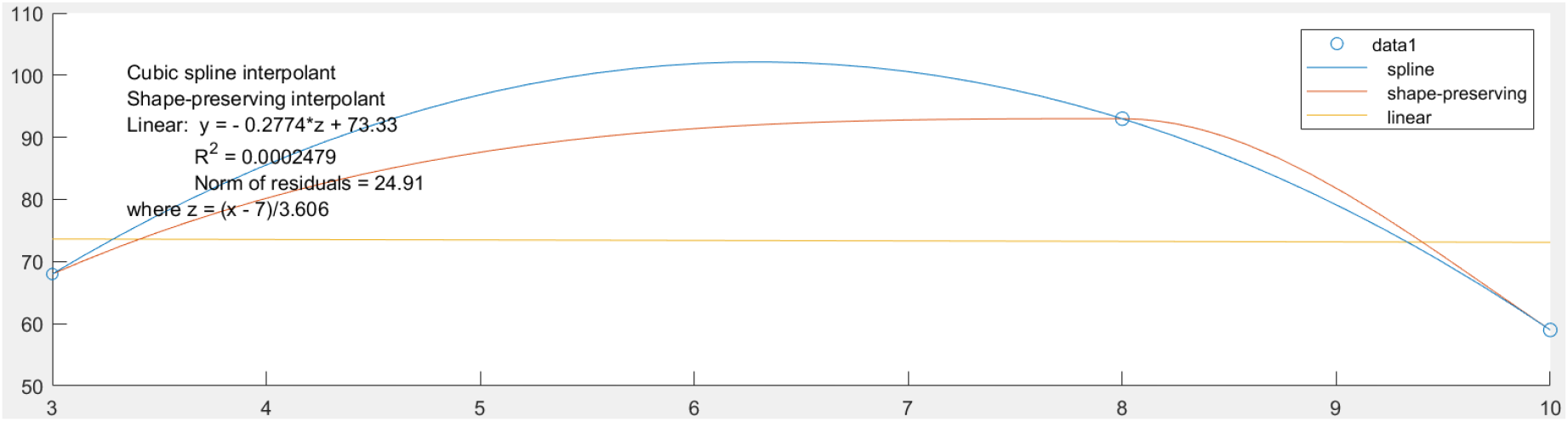
The Effect Size of Ejection Fraction Value < 50% on Mean Arterial Pressure. In figure 3, the x-axis shows the total dose of propofol in mg/ml given by the anesthesiologist and the y-axis shows the amount of MAP produced after induction of propofol. The number of subjects in this group were 3 patients, with the results of p-value = 0.97. In this case, the researchers observed the magnitude of changes in MAP resulting from the induction of propofol to patients. Based on the results of data analysis, the induction dose of propofol given to patients with ejection fraction value <50% did not cause any changes in MAP even though at that dose of propofol could significantly change the BIS index value.

**Figure 4.**
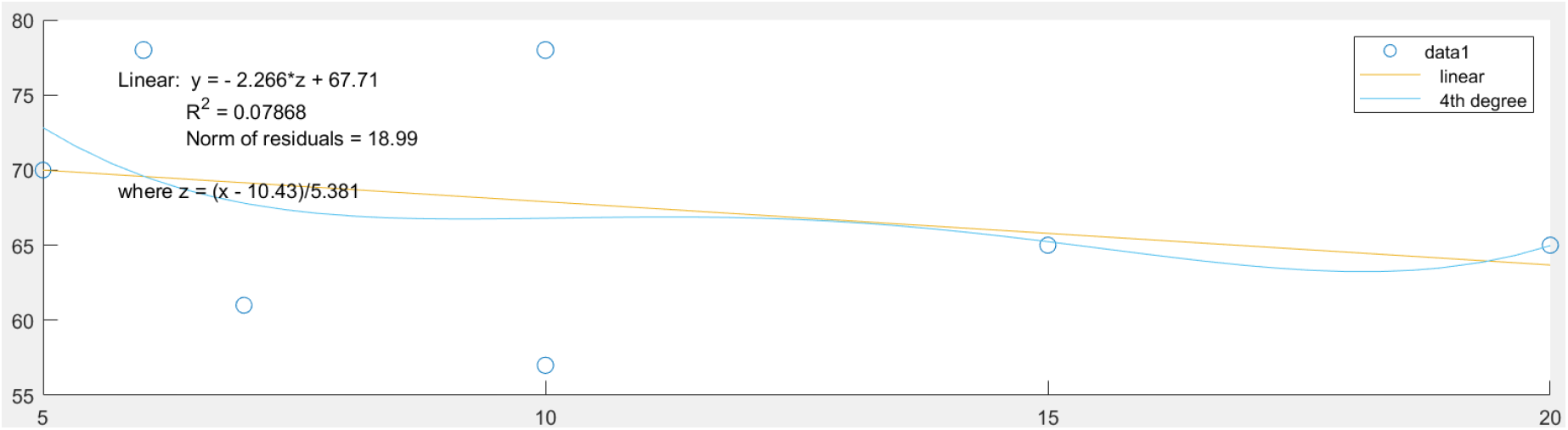
The Effect Size of Ejection Fraction Value ≥ 50% on Mean Arterial Pressure. In figure 4, the x-axis shows the number of doses of propofol in mg/ml given by the anesthetist and the y-axis shows the amount of MAP produced after the induction of propofol. The number of subjects included in this group were 6 patients, with the results of the analysis p-value = 0.005. Based on the results of data analysis using multivariate linear regression in the group of patients with ejection fraction value of ≥ 50%, the induction dose of propofol can produce a small effect on changes in patients’ MAP.

**Figure 5.**
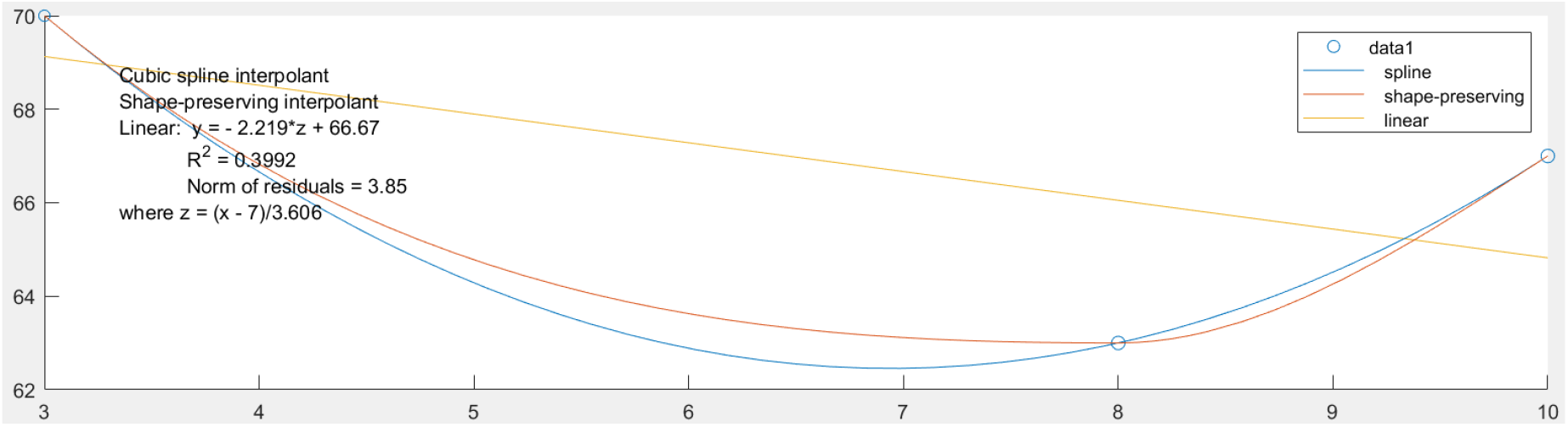
The Effect Size of Ejection Fraction Value < 50% on Heart Rate. In figure 5, the x-axis shows the dose of propofol given as anesthetic induction and the y-axis shows the heart rate response resulting after propofol administration. The number of subjects included in this group were 3 patients, with the results of p-value = 0.06. Based on this figure, the induction dose of propofol given to the patient group with ejection fraction value of <50% produced a moderate effect on the magnitude of change in heart rate, this can be seen in the R^2^.

**Figure 6.**
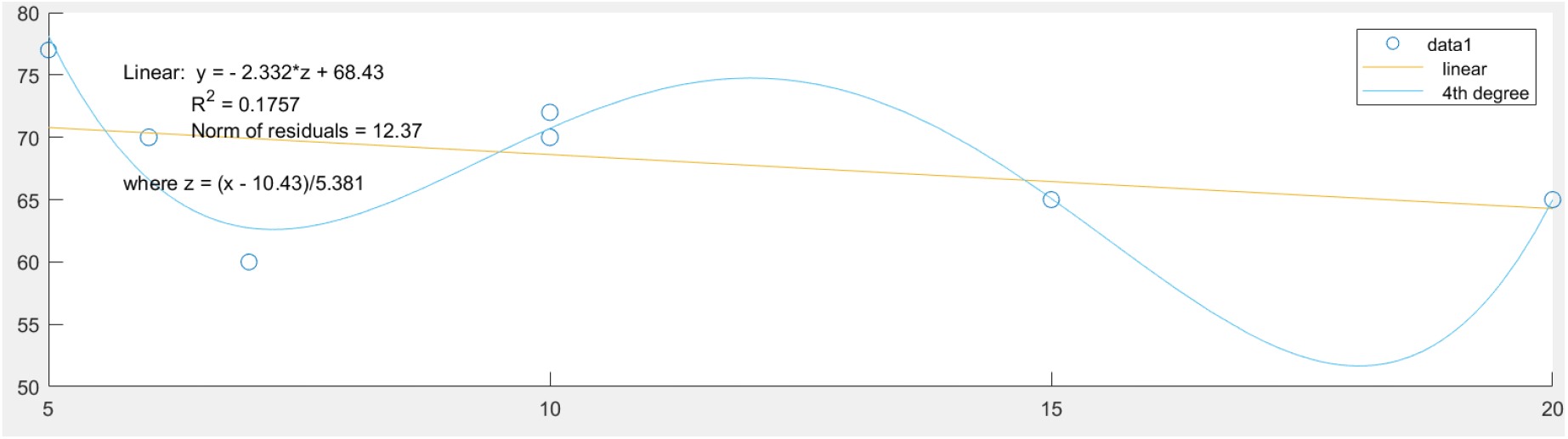
The Effect Size of Ejection Fraction Value ≥ 50% on Heart Rate. In figure 6, the x-axis shows the amount of propofol given as an anesthetic induction dose in mg/ml and the y-axis shows the change in heart rate resulting after propofol induction. The number of subjects included in this group were 6 patients, with the results of p-value = 7.0776e-04. The induction of propofol in the group of patients with ejection fraction value of ≥ 50% had a very small effect on heart rate, although the induction dose of propofol was able to change the BIS index value.

## DISCUSSION

The use of propofol as an anesthetic induction agent in general anesthesia is a good choice for most operations, however, the use of propofol has several criteria for the patient’s physiological condition that must be met to minimize the side effects that occur due to the use of propofol ^(1)^. Especially in patients undergoing cardiac surgery, the use of propofol as an anesthetic induction agent requires several titrations at the time of administration which ideally is 40mg/ml every 10 seconds for each titration, carried out until the onset of propofol begins. This attempt is aimed to minimize the side effects that occur as a result of administering propofol ^(14)^.

In CABG patients, propofol as an anesthetic induction agent causes a change in the BIS index value according to the expected range of 40-60 by administering a smaller induction dose of propofol than the usual induction dose to patient non-cardiac surgery. Based on the results of the data analysis of this observational study, at this dose of propofol had almost no effect on the patient’s MAP or heart rate where in the general use of propofol would cause a dramatic decrease in MAP depending on the administering dose of propofol. The decrease in MAP that occurs due to the administration of propofol is generally temporary, however, the excessive doses of propofol cause depression in the airways and myocardium ^(15–16)^, and the risk of cardiac arrest which can cause sudden death is increased in patients with ASA status III or more ^(17)^.

Based on the data of this observational study, certain induction doses of propofol which are relatively smaller than usual for patients with EF values <50% who will undergo CABG can produce BIS index values that are in line with the expectations of anesthesiologists without causing a significant decrease in MAP, and the propofol induction doses which are relatively greater in this group of patients can cause a significant decrease in BIS index value, which is less than 40, and it may result in postoperative cognitive dysfunction or known as delayed neurocognitive recovery ^(18)^. Likewise in the group of patients with ejection fraction value of ≥ 50%, the induction dose of propofol required to produce BIS index value that corresponds to the range of 40-60 is greater when compared to the group of patients with ejection fraction value of <50%. In CABG patients with ejection fraction < 30%, anesthetists no longer use propofol as an induction agent in general anesthesia so that these patients were excluded from being involved in this study.

This observational study used multivariate linear regression analysis to determine the effect size of ejection fraction value, which in this study was indicated by the R squared (R^2^) on the propofol induction doses requirement and several outcomes, namely the BIS index, MAP, and heart rate values respectively in patients who are about to undergo CABG surgery. The R^2^ value has several classifications to determine the magnitude of influence or the effect size produced by a variable. Some of the R^2^ classifications are small (0.10 - < 0.30), medium (0.30 - < 0.50), and large (≥ 0.50) ^(19)^. In addition to the R^2^ value, this observational study also produced p-values for each different condition to determine the statistical significance of each of these conditions. However, the p-value is greatly influenced by sample size and the strength of a relationship along those variables ^(20)^, so that in addition to the p-value, the effect size or influence of a variable is much more important for a study to be considered in determining conclusions ^(21)^. The primary outcome in this observational study is the BIS index value where an anesthetist can determine the adequacy of anesthetic to produce unconsciousness without causing a coma (overdose of anesthetic) or awareness (inadequate anesthesia). The method of determining the level of consciousness of anesthetized patients using the BIS spectral index is an effective method based on a systematic review and meta-analysis study ^(22)^, it was a result of BIS spectral index has a very good sensitivity in detecting brain waves such as alpha, beta, theta, delta, and gamma where in general hypnotic anesthetic drugs will produce delta waves ^(23)^. In addition, the BIS index method has also received approval from the FDA to determine the level of consciousness in patients and has excellent sensitivity to propofol when compared to midazolam and sevoflurane ^(10)^.

Thus, based on the primary outcome of this observational study, namely the BIS index value, the classification of patients who will undergo CABG surgery according to their ejection fraction value is an appropriate method to minimize moderate risks due to the use of propofol such as a dramatic decrease in MAP and awareness during surgery, to severe risks such as CNS depression in cardiac surgery patients receiving propofol induction.

### Limitations Study

In this study, the number of research subjects who met the inclusion criteria was nine patients of which the number consisted of 6 patients with normal ejection values and 3 patients with a reduced ejection fraction. So that some of the outcomes observed in this study have p-values that are not statistically significant even though they have a strong impact based on the changes produced.

## CONCLUSIONS

In CABG patients with a mid-reduced ejection fraction value, the administration of propofol as an induction in general anesthesia can have a huge influence on the changes in BIS index value when compared to the group of patients with normal ejection fraction values based on the R^2^ generated by multivariate linear regression analysis using MATLAB R2022a software.

## Data Availability

All data produced in the present work are contained in the manuscript

## AUTHOR CONTRIBUTIONS

Conceptualization, data analysis, and software : first author

Supervision : all the supervisors from Clinical Pharmacy Department, Cardiac Anesthesiology Department, and Cardiothoracic and Vascular Surgery Department

Methodology : first author Writing original draft : first author Review and editing : first author

## DATA AVAILABILITY STATEMENT

All data produced in the present work are contained in the manuscript

## ACKNOWLEDGMENTS

The author would like to thank the Supervisor of the Clinical Pharmacy Department at Universitas Airlangga, Surabaya, Indonesia, for supervising the author, all Cardiac Anesthesiologist in the Department of Cardiac Anesthesiology at Dr.Soetomo General Hospital and Premier Hospital in Surabaya, Indonesia, who are involved in the surgery and giving advice, and the Cardiothoracic and Vascular Surgeon in the Department of Cardiothoracic and Vascular Surgery at Dr. Soetomo General Hospital and Premier Hospital in Surabaya, Indonesia, for supervising the author and involved in the surgery.

